# Brain region- and solubility-dependent proteomic signatures of neurofibrillary tangle pathology in Alzheimer’s disease

**DOI:** 10.64898/2026.10.04.26361149

**Authors:** Xuemei Zeng, Eric E. Abrahamson, Wasiu G. Balogun, Julia Kofler, William R. Paljug, Violetta N. Pivtoraiko, Milos D. Ikonomovic, Thomas K. Karikari

## Abstract

Neurofibrillary tangles (NFTs) containing hyperphosphorylated tau proteins are a hallmark neuropathology of Alzheimer’s disease. However, the molecular basis underlying their spatial heterogeneity and biochemical complexity remain unclear. To address this, we performed mass spectrometry–based proteomic profiling of two neocortical regions with different susceptibilities to NFT pathology—the inferior temporal (IT) and middle frontal (MF) cortices—from 27 autopsy cases stratified by Braak NFT stage (Low 0–II, n=8; Mid III–IV, n=11; High V–VI, n=8). Brain homogenates were sequentially fractionated using buffers of increasing solubility (TBS, Na₂CO₃, and Urea), followed by trypsin digestion and analysis by nanoLC–MS/MS with data-independent acquisition. Proteins were identified using PEAKS Studio 12 and linear mixed-effects models were applied to assess Braak stage–associated and region-specific changes in protein abundance and solubility. A total of 5,573 protein groups were identified, including 452 differentially expressed proteins (DEPs), 183 bidirectional expression proteins (BEPs), and 693 differentially soluble proteins (DSPs) associated with NFT pathology. These proteins were enriched in pathways related to cytoskeletal organization, synaptic transmission, metabolism, vesicular trafficking, and proteostasis. Marked regional differences were observed, with limited overlap between IT and MF across DEPs, BEPs, and DSPs. Protein change in MF was dynamic with a higher number of BEPs and suggested active cytoskeletal remodeling, whereas IT showed fewer BEPs and evidence of structural disruption, including alterations in intermediate filament components, consistent with vulnerability of IT and possible compensatory/plasticity changes in MF cortex. MF cortex had the greatest downregulation of DEPs between Low and Mid Braak stages, while the transition from Mid to High Braak stage showed upregulation of most of DEPs in both regions. In addition, widespread changes in protein solubility were observed, particularly decreased solubility at later Braak stages, implicating disruption of proteostasis. Together, these findings highlight region- and stage-dependent proteomic alterations associated with tau pathology and underscore their relevance for understanding disease mechanisms and informing biomarker development.

## Background

Alzheimer’s disease (AD) is a neurodegenerative disorder characterized neuropathologically by brain accumulation of fibrillar aggregates including amyloid-beta (Aβ) peptides in extracellular plaque deposits and over-phosphorylated microtubule-associated protein tau (p-tau) in intracellular neurofibrillary tangles (NFT) consisting of [2, 15]. While relative contribution of these pathologies to clinical symptoms remains unclear, the NFT pathology is more closely associated with cognitive impairment than Aβ plaques [38]. Furthermore, anti-Aβ therapies have minimal or only modest impact on cognitive decline [1, 13, 39], suggesting that tau pathology may play a key role in cellular degeneration, synaptic impairment, and neural network dysfunction characterizing the disease. However, clinical trials targeting tau pathology have had limited success. This could be due, in part, to knowledge gaps related to the molecular mechanisms of tau pathobiology [12, 45]. In AD, cortical NFT initially develop in the mesial temporal lobe transentorhinal and entorhinal regions before they emerge in the hippocampus and subsequently in neocortical association cortices, a pattern established as six Braak NFT stages [10]. The hierarchical Braak staging scheme allows for investigations into brain areas that are affected with NFT tau pathology at different time points during the progression of AD.

Recently, proteomics has become a cornerstone for elucidating the molecular and cellular mechanisms underlying AD pathobiology [17, 42, 55]. Emerging technologies, including mass spectrometry (MS)-based approaches now allow deep proteomic profiling of thousands of proteins simultaneously [22, 40]. Advances in MS-based techniques, including enhanced sample preparation, greater instrument sensitivity, faster scan speeds, optimized data acquisition strategies, and improved bioinformatics, have significantly expanded proteome coverage and improved quantification accuracy [22, 23, 35, 51]. MS offers unbiased, high-resolution protein profiling through direct peptide sequencing. This capability provides unmatched specificity, enabling precise identification of proteins, isoforms, and post-translational modifications. The utility of MS-based proteomics has been demonstrated in numerous studies and highlighted in recent reviews emphasizing its potential to uncover key molecular mechanisms driving AD pathology [18, 27, 30, 42, 54].

Despite these advances, the spatial heterogeneity and biochemical complexity of tau NFT pathology remain incompletely understood. Most proteomic studies of AD brain have focused on a single brain region, with frontal cortex and hippocampus analyzed most frequently [4, 30]. Furthermore, the majority of studies focused on unfractionated whole-tissue homogenates (homogenates of a single solubility type) to identify differences in protein abundance. In the current study, we hypothesized that proteomics resulting from biochemical fractionation of brain tissue samples, which use different extraction buffers to separate proteins according to their solubility, provide a more refined insight into protein abundance changes associated with tau aggregation in its diffusible, membrane bound, or filamentous aggregation state. To test this hypothesis, we conducted MS-based quantitative proteomic analyses of two neocortical regions with distinct vulnerability to NFT pathology, the inferior temporal (IT) and middle frontal (MF) cortex. The IT cortex is among the earliest neocortical areas affected by NFT (Braak NFT stage III-IV), whereas frontal cortex becomes involved at later stages (NFT Braak stage V-VI) and exhibits evidence of synaptic and functional plasticity in prodromal AD [6, 11, 14, 26, 36]. By assaying these brain regions, we sought to integrate protein abundance and solubility profiling to capture molecular alterations spanning different phases of NFT pathology across early, mid and late Braak NFT Stages, to provide a more comprehensive view of AD proteinopathy during tau pathobiology progression.

## Methods

### The autopsy cohort

Study was approved by the Committee for Oversight of Research and Clinical Training Involving Decedents (CORID) and appropriate institutional review boards at the University of Pittsburgh, in accordance with the Declaration of Helsinki. Postmortem frozen brain tissue samples were obtained from the brain bank in the Alzheimer’s Disease Research Center (ADRC) at the University of Pittsburgh. The autopsy cohort (n=27) involved eight cases with low to no NFT burden (Braak NFT Stages 0-II), 11 cases with intermediate (mid) NFT burden (Braak NFT Stages III-IV), and eight cases with high level of NFT pathology (Braak NFT Stages V-VI). Demographic and neuropathological characteristics of the cases are provided in Table 1. The median age at death among cases was 82 years (interquartile range [IQR]: 16 years), with 52% being female. Neuropathology evaluation included standard diagnostic neuropathology procedures utilizing immunohistochemical analyses of Aβ, phosphorylated tau, α-synuclein, and transactive response DNA-binding protein (TDP-43), as well as routine hematoxylin and eosin and Bielschowsky silver stain [37].

**Table 1:**
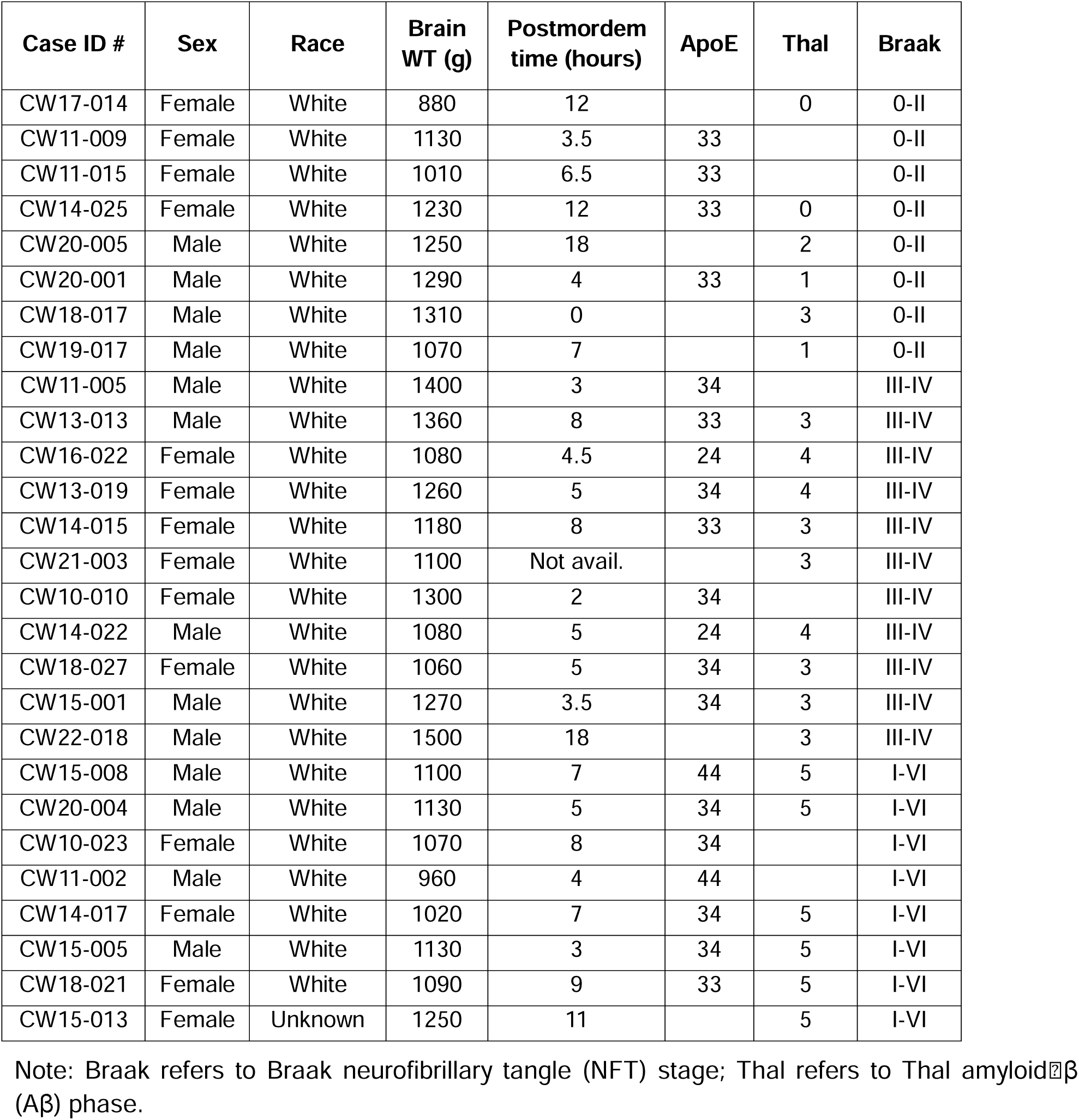
Demographic and neuropathological characteristics of the study cohort.

| Case ID # | Sex | Race | Brain WT (g) | Postmordem time (hours) | ApoE | Thal | Braak |
| --- | --- | --- | --- | --- | --- | --- | --- |
| CW17-014 | Female | White | 880 | 12 |  | 0 | 0-II |
| CW11-009 | Female | White | 1130 | 3.5 | 33 |  | 0-II |
| CW11-015 | Female | White | 1010 | 6.5 | 33 |  | 0-II |
| CW14-025 | Female | White | 1230 | 12 | 33 | 0 | 0-II |
| CW20-005 | Male | White | 1250 | 18 |  | 2 | 0-II |
| CW20-001 | Male | White | 1290 | 4 | 33 | 1 | 0-II |
| CW18-017 | Male | White | 1310 | 0 |  | 3 | 0-II |
| CW19-017 | Male | White | 1070 | 7 |  | 1 | 0-II |
| CW11-005 | Male | White | 1400 | 3 | 34 |  | III-IV |
| CW13-013 | Male | White | 1360 | 8 | 33 | 3 | III-IV |
| CW16-022 | Female | White | 1080 | 4.5 | 24 | 4 | III-IV |
| CW13-019 | Female | White | 1260 | 5 | 34 | 4 | III-IV |
| CW14-015 | Female | White | 1180 | 8 | 33 | 3 | III-IV |
| CW21-003 | Female | White | 1100 | Not avail. |  | 3 | III-IV |
| CW10-010 | Female | White | 1300 | 2 | 34 |  | III-IV |
| CW14-022 | Male | White | 1080 | 5 | 24 | 4 | III-IV |
| CW18-027 | Female | White | 1060 | 5 | 34 | 3 | III-IV |
| CW15-001 | Male | White | 1270 | 3.5 | 34 | 3 | III-IV |
| CW22-018 | Male | White | 1500 | 18 |  | 3 | III-IV |
| CW15-008 | Male | White | 1100 | 7 | 44 | 5 | I-VI |
| CW20-004 | Male | White | 1130 | 5 | 34 | 5 | I-VI |
| CW10-023 | Female | White | 1070 | 8 | 34 |  | I-VI |
| CW11-002 | Male | White | 960 | 4 | 44 |  | I-VI |
| CW14-017 | Female | White | 1020 | 7 | 34 | 5 | I-VI |
| CW15-005 | Male | White | 1130 | 3 | 34 | 5 | I-VI |
| CW18-021 | Female | White | 1090 | 9 | 33 | 5 | I-VI |
| CW15-013 | Female | Unknown | 1250 | 11 |  | 5 | I-VI |

Note: Braak refers to Braak neurofibrillary tangle (NFT) stage; Thal refers to Thal amyloid $\beta$ (Aβ ) phase.

### Overview of the experimental workflow

Supplementary Fig. S1a illustrates the experimental workflow used in this study. In brief, high-resolution liquid chromatography tandem mass spectrometry (LC-MS/MS) analysis was performed on 160 fractionated tissue homogenates of postmortem frozen IT cortex (Brodmann area 20) and MF cortex (Brodmann area 46) from cases at different severity of Braak NFT stages (Low, Mid and High). Each brain sample was sequentially fractionated into three components using different extraction buffers: TBS (enriching diffusible proteins), Na_2_CO_3_ (enriching membrane-associated proteins), and urea (enriching fibrillar proteins). Two MF samples from Mid-Braak donors, one Na_2_CO_3_ fraction and one Urea fraction, were lost during preparation. The final dataset included 81 IT samples and 79 MF samples (Supplementary Fig. S1b).

### Postmortem brain sample processing and tissue fractionation

Samples of the IT cortex and MF cortex were dissected from each brain at autopsy, immediately frozen on dry ice, and stored at -80 °C until use. Samples were homogenized using a glass Dounce homogenizer at a concentration of 300 mg/ml in Tris buffered saline (TBS; 50mM Tris pH 8.0, 100mM NaCl) with protease inhibitors (PI; AEBSF, 40X dilution; Sigma Protease Inhibitor Cocktail P8340, 100X dilution; Sigma Phosphatase Inhibitor Cocktail 2 P5726, 100X dilution; Sigma Phosphatase Inhibitor Cocktail P0044, 100X dilution) and sonicated on ice for ten seconds. Samples were centrifuged at 135,000×g for 30 minutes at 4 ⁰C, and the supernatants were collected as the <u>TBS-soluble fraction</u>. The pellet was then resuspended in the same volume as centrifuged in the previous step, using ice cold 100 mM Na_2_CO_3_ (pH 11.0) with protease inhibitors, sonicated for ten seconds on ice, and incubated on ice for 20 minutes. This sample was centrifuged at 135,000×g for 30 minutes at 4 ⁰C. The supernatant was saved as the <u>Na_2_CO_3_-soluble fraction</u>. The pellet was then resuspended in the same volume as centrifuged in the original sample homogenate, using urea buffer (7 M urea, 2 M thiourea, 4% CHAPS, 30 mM bicine, pH 8.5), sonicated on ice for ten seconds, and incubated on ice for 20 minutes. The sample was then centrifuged using a bench-top, refrigerated microcentrifuge at 135,000 x g for 30 minutes at 4 ⁰C, and the supernatant was carefully removed as the <u>Urea-soluble fraction</u> and kept on ice. All soluble and fibrillar fractions were aliquoted at time of collection and stored at -80°C until assayed.

### Proteomic sample preparation

For each preparation, 30 μg of the brain lysate was processed using the single-pot, solid-phase-enhanced sample-preparation (SP3) on-bead digestion protocol with trypsin as described previously [20]. In brief, brain lysate was adjusted to a final volume of 200 μL using 100 mM Tris, pH 8.0/2% sodium dodecyl sulfate (SDS) and then subjected to reduction with 10 mM dithiothreitol (DTT) and alkylation with 20 mM iodoacetamide (IAA). Subsequently, 1000 μL of 100% ethanol and 30 μL of 20 mg/mL pre-washed SP3 paramagnetic bead mix (an equal mix of hydrophobic beads [GE Healthcare Bio-Sciences Cat #65152105050250] and hydrophilic beads [GE Healthcare Bio-Sciences Cat #45152105050250]) were added, followed by a 20-minute incubation with vigorous shaking (1400 rpm). The beads were then washed three times with 80% ethanol, all liquid was removed, resuspended in 100 μL of 50 mM ammonium bicarbonate/1 mM CaCl₂ containing 1 μg of trypsin, and incubated overnight at 37°C with vigorous shaking (1400 rpm). The supernatant, containing the resulting tryptic peptides, was transferred to a new tube. To ensure complete recovery of peptides, 100 μL of 0.1% trifluoroacetic acid (TFA)/2% acetonitrile (ACN) was added to wash the SP3 beads, and the wash was combined with the previous supernatant. Tryptic peptides were desalted using C18 solid-phase extraction (SPE) cartridges (Nest Group, BioPureSPN Mini), speed-vac dried, and resuspended in 60 μL 0.1 formic acid (FA).

### Liquid chromatography tandem mass spectrometry (LC-MS/MS)

Peptides were analyzed using nanoscale LC-MS/MS as described previously [20]. For each analysis, 1 μL of the peptides were loaded using a nanoflow HPLC system (Dionex UltiMate 3000 RSLCnano UPLC System, ThermoFisher Scientific) onto an Aurora 5 cm x 150 µm C18 electrospray ionization column (Ionopticks, #AUR2-50150C18A). The HPLC system was online coupled to an Orbitrap Exploris™ 480 mass spectrometer (ThermoFisher Scientific). For unbiased proteomics, peptides were eluted from the column over a 65-minute gradient at a flow rate of 1 µL/min using 0.1% formic acid in water as mobile phase A and 0.1% formic acid in acetonitrile as mobile phase B. The gradient was set as follows: 0-0.5 min, 3% B; 0.5-60.5 min, linear increase to 30% B; 60.5-61 min, increase to 90% B; maintained at 90% B until 62.1 min; 62.1-62.5 min, decrease to 3% B; 62.5-65 min, held at 3% B. The mass spectrometer was operated in positive ionization mode with an electrospray voltage of 1.2 kV and a capillary temperature of 275 °C. MS data were acquired using a data-independent acquisition (DIA) method with 3 second cycle time, consisting of one full MS1 scan followed by 50 isolation windows (width: 11 m/z, overlap: 1 m/z) covering the range 349.5–910.5 m/z, plus a final window spanning 909.5–1400 m/z. Full-scan MS1 spectra were collected at a resolution of 45,000 with a scan range of 350–1400 m/z, a normalized automatic gain control (AGC) target of 300% and a maximum injection time of 20 milliseconds. MS2 spectra were acquired at a resolution of 30,000 using a normalized AGC target of 200%, a maximum injection time of 35 milliseconds and a normalized collision energy (NCE) of 30%. RF lens were set at 45% for full MS and 50% for MS2 scans.

Aβ42 was quantified by parallel reaction monitoring (PRM) using a 10-min chromatographic gradient consisting of 3-34% solvent B over 5.5 min, followed by 90% solvent B for 1 min and re-equilibration at 3% solvent B for 3.5 min at a flow rate of 1 μL/min. Orbitrap MS/MS spectra were acquired at a resolution of 15,000. The precursor ion (m/z 633.384) was isolated using a 2 m/z window and fragmented by higher-energy collisional dissociation (HCD) with a normalized collision energy of 30%. Aβ42 abundance was quantified using the summed peak areas of the y2, b5, b6, b8, and b10 fragment ions.

### MS data processing

Raw mass spectrometry data were analyzed using PEAKS Studio 12 for peptide/protein identification and label-free quantification. A library-free, direct database search workflow was applied to match DIA spectra against the Human SwissProt database (downloaded on December 12, 2023), which contains 17,190 entries. The precursor mass tolerance was set to 20 ppm and fragment mass tolerance to 0.02 Da. Carbamidomethylation (+57.02 Da) was specified as a fixed modification, while methionine oxidation (+15.99 Da) was set as a variable modification. Maximum one variable post-translational modification (PTMs) per peptide was allowed. Trypsin was selected as the digestion enzyme, permitting up to two missed cleavages and restricting peptide lengths to 7-30 amino acids. Results were filtered at a peptide-level false discovery rate (FDR) of 1.0%, followed by a protein group-level FDR of 1.0%, requiring at least one unique peptide per protein. Protein abundances were normalized to total protein area prior to downstream statistical analysis. For simplicity, proteins are referenced by gene symbols throughout figures and tables to align with mass spectrometry-based protein identification output.

### Statistical and bioinformatic analyses

Statistical analyses were conducted using MATLAB (R2024a). Prior to analysis, protein areas were normalized to the total protein area of their respective samples. Log₂ transformation was applied to the normalized values to improve data normality.

To identify proteins showing consistent Braak stage-associated changes across extraction fractions, differential expression analyses were performed using linear mixed-effects models implemented in MATLAB (fitlme), separately for each brain area (IT and MF), with log₂-transformed normalized protein area as the dependent variable. The models were fit separately for each protein using the formula: Protein area ∼ Braak group + Fraction + (1|Participant ID). NFT Braak groups (Low-, Mid-, and High-Braak) and fraction types (TBS, Na_2_CO_3_, and Urea) were included as categorical fixed predictors, and a random intercept was added to account for brain donor-specific effects. Differentially expressed proteins (DEPs) were defined based on predefined pairwise Braak-stage contrasts (Mid-Braak vs Low-Braak, High-Braak vs Low-Braak, and High-Braak vs Mid-Braak). Effect sizes were estimated from model coefficients, and significance was assessed using model-derived contrast tests. A protein was considered differentially expressed if at least one pairwise Braak-stage comparison met the criteria of p < 0.005 and an absolute effect size (|β|) > 0.3.

To evaluate potential bidirectional changes in abundance across Braak stages, Braak group was included in the linear mixed models as an ordinal variable along with its quadratic term: Protein abundance ∼ Braak group + Braak group² + Fraction + (1|Participant ID). Bidirectionally expressed proteins (BEPs) were defined as proteins exhibiting a significant quadratic effect (p < 0.005 and |β| > 0.3), together with significant pairwise differences between Mid-Braak and Low-Braak groups and between High-Braak and Mid-Braak groups (p < 0.05 for both comparisons). To identify differentially soluble proteins (DSPs), fraction type was modeled as an ordinal variable, assuming decreasing solubility across TBS, Na_2_CO_3_, and Urea (coded as 1, 2, and 3, respectively). A linear mixed-effects model was fitted as follows: Protein abundance ∼1+Braak*Fraction+(1|Participant ID).

Given the exploratory nature of the study, no multiple testing correction was applied for all statistical tests. Instead, a more stringent p-value threshold (p < 0.005) was used in combination with an effect size criterion (|β| > 0.3) for each pairwise comparison to prioritize biologically meaningful signals while reducing the likelihood of identifying changes with small effect sizes. To identify concordant NFT Braak stage associated proteomic changes, we used fraction-adjusted correlation to evaluate the consistency between tissues. Protein abundances were first adjusted for solubility fraction by regressing out fraction effects within each tissue using linear mixed models: Protein abundance ∼ fraction + (1|Participant ID). Spearman correlations were then performed on the resulting residuals to quantify inter-regional concordance independent of fraction-specific abundance differences. A coefficient > 0.6 was considered strong correlation.

Several online resources were utilized for the bioinformatic analysis, including WebGestalt [19, 57], David Bioinformatic Tool [25, 46], Metascape [58], and String-db [47, 49]. Visualization of bioinformatic results was performed using SRplot and Cytoscape [44, 50].

## Results

### Overview of LC-MS/MS results

A total of 5,746 protein groups were identified from the combined analysis of 160 fractionated homogenates derived from frozen IT and MF tissues (n = 54) across 27 cases (Supplementary Fig. S1b). The number of protein groups and peptide sequences ranged from 4,385 to 5,582 and 37,478 to 78,069, respectively, for each individual homogenate, with Urea fractions yielding the highest numbers of proteins and peptides, followed by Na₂CO₃ and then TBS fractions (Supplementary Fig. S1c–d).

To ensure high identification confidence, we filtered the results to include only protein groups with at least two unique peptides, yielding a final set of 5,573 proteins that were quantified by normalized protein area (See Methods) across all samples (Supplementary Table S1). Gene Ontology analysis of these proteins revealed significant functional overrepresentation of proteins associated with synapse organization (n=292), vesicle-mediated transport in synapse (n=177), ribose phosphate metabolism (n=302), cytoplasmic translation (n=132), and Golgi vesicle transport (n=205) (Fig. 1a). In terms of cellular components, the proteins were enriched in the glutamatergic synapse (n=302), neuron-to-neuron synapses (n=264), the postsynaptic specialization (n=251), and the mitochondrial matrix (n=316). For molecular functions, enrichment was observed in proteins involved in cadherin binding (n=252), actin binding (n=244), and GTPase activity (n=190). KEGG enrichment analysis using ShinyGO identified many proteins previously implicated in human neurodegenerative diseases, including AD, Prion disease, Parkinson’s disease, and amyotrophic lateral sclerosis (Fig. 1b), supporting its utility for investigating disease-relevant molecular pathways.

**Fig. 1.**
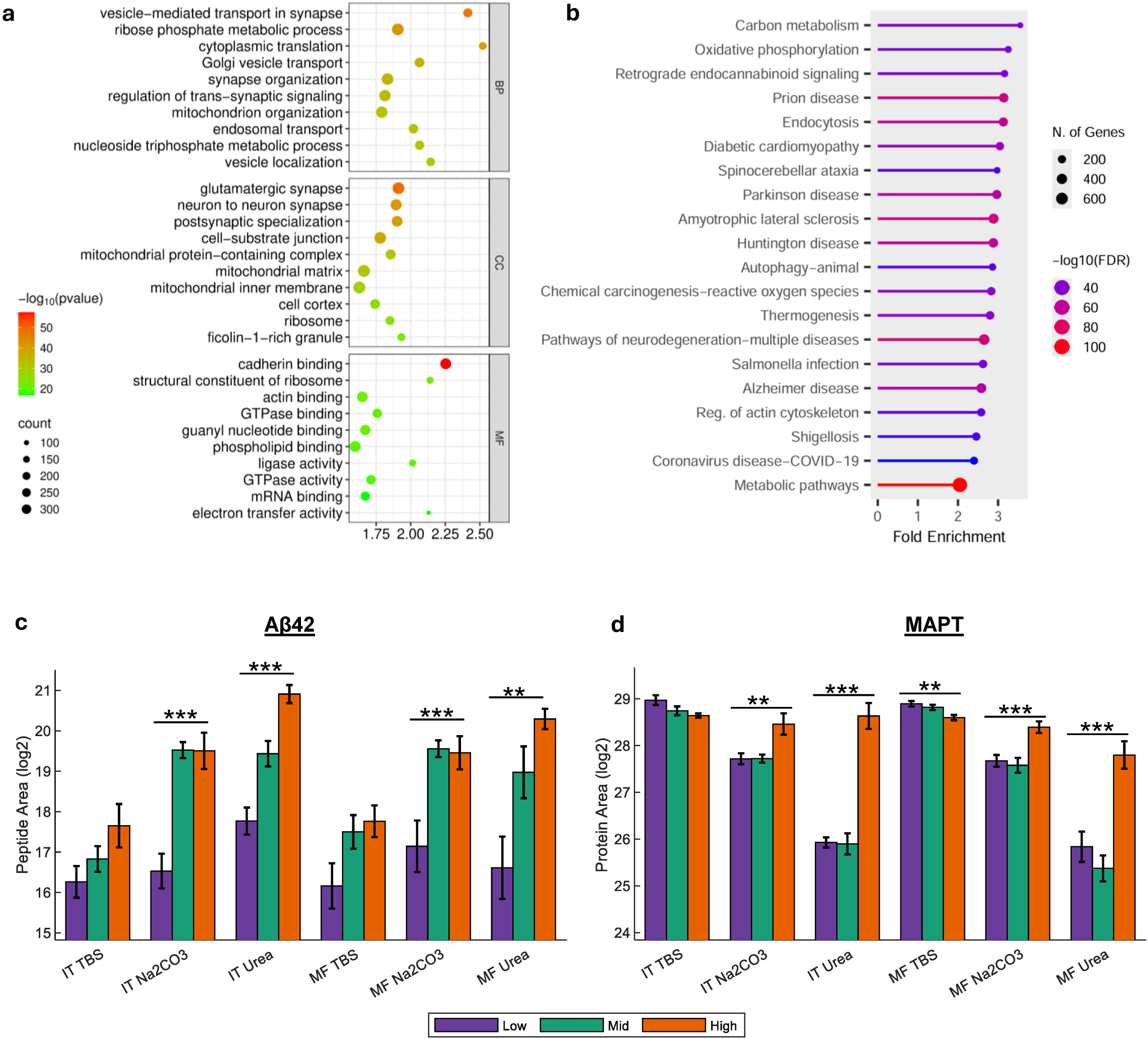
Overview of LC-MS/MS results. (**a)** Gene Ontology (GO) enrichment analysis of all identified proteins. The x-axis represents the enrichment score. BP, biological process; MF, molecular function; CC, cellular component. (**b**) Enrichment of KEGG pathways by ShinyGO. (**c-d**) Bar plots showing log₂-transformed peak areas of Aβ42 (**c**) and MAPT (**d**), stratified by NFT Braak group. Aβ was quantified by peptide GAIIGLMVGGVVIA. Error bars represent the standard error of the mean. Low, Mid, and High correspond to NFT Braak stages 0–II, III–IV, and V–VI, respectively. * p < 0.05; ** p < 0.01; *** p < 0.001. Point color reflects statistical significance, and point size indicates the number of proteins in each category for panels a and b.

As expected, primary components of the two pathological hallmarks of AD, MAPT (microtubule-associated protein tau), and Aβ42 (quantified using the Aβ42-specific C-terminal peptide GAIIGLMVGGVVIA), the predominant Aβ species deposited in amyloid plaques, exhibited significant abundance differences related to Braak NFT stages (Fig. 1c-d). Greater abundance of neocortical Aβ levels became evident at the Mid-Braak stage compared to Low-Braak, with significant difference observed in the Na_2_CO_3_ (both regions, p < 0.001) and Urea fractions (IT: p<0.001; MF, p=0.002). Although Aβ42 showed a similar increasing trend in the TBS fraction, the differences did not reach statistical significance. In the Na_2_CO_3_ fraction, Aβ42 abundance plateaued at the Mid-Braak stage, whereas it continued to increase in the Urea fraction with advancing Braak stage. In contrast, MAPT abundance showed a decreasing trend in the TBS in both regions, reaching significant in MF (p=0.006) but not in IT, while exhibiting significant increases in both Na_2_CO_3_ and Urea fractions. Unlike Aβ42, the increases in MAPT became evident only at the High-Braak stage, with the largest differences observed in the Urea fraction in both regions.

### Identification of differentially expressed proteins (DEPs) by Braak stage and by region

We applied linear mixed-effects models to identify DEPs changes across the spectrum of NFT pathology progression from low (stage 0-II) to mid (stage III, IV) to high (stage V, VI) Braak NFT groups, stratified by tissue region. A protein was classified as a DEP if it met the significance threshold (p < 0.005) in at least one of the pairwise comparisons among the Braak NFT groups and had an absolute beta coefficient (|β|) >0.3. Fig. 2a–2f display the -log₁₀ p-values for the main effect of Braak NFT group alongside the corresponding β estimates (z-normalized) from each pairwise comparison.

**Fig. 2.**
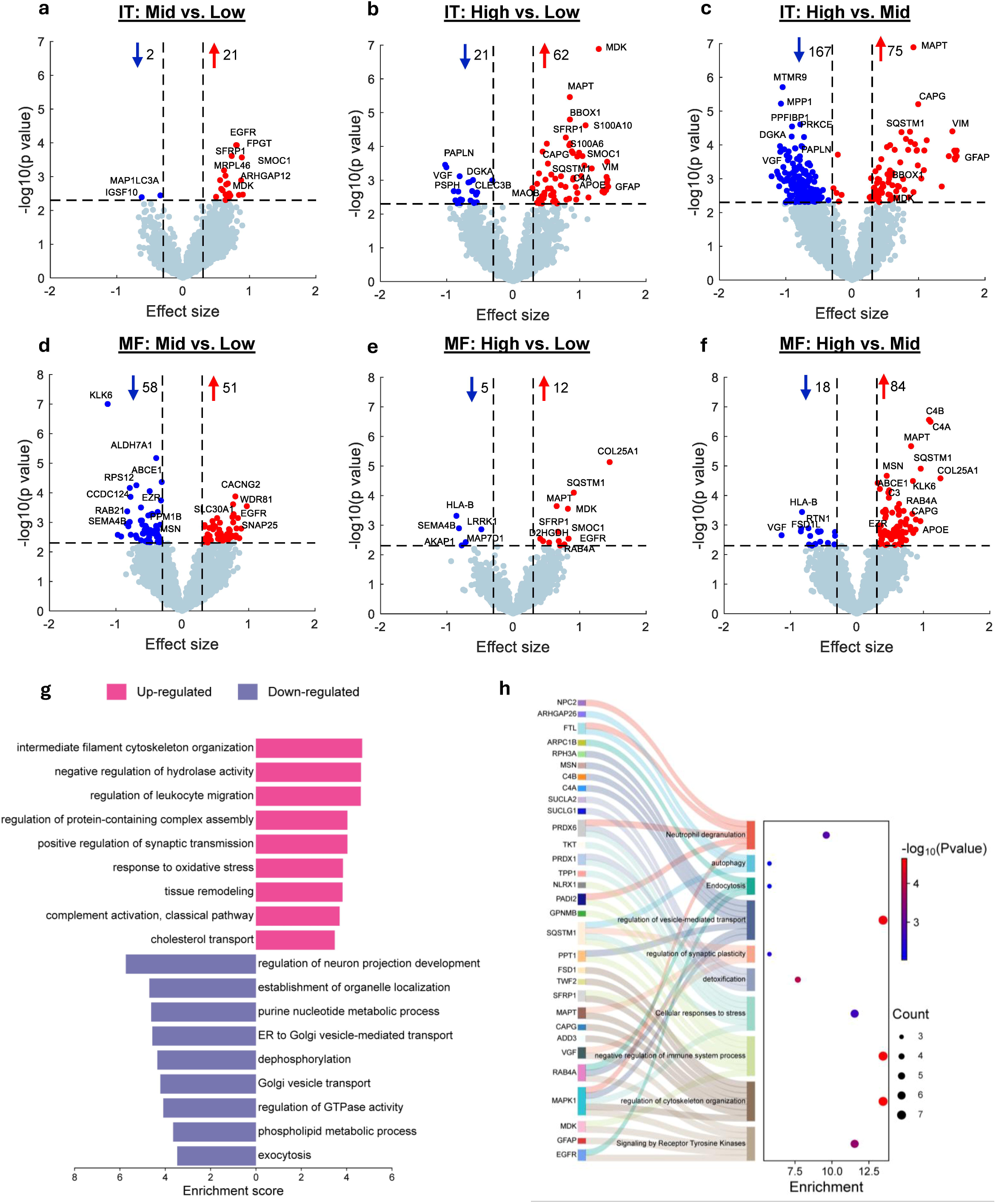
Identification of proteins with NFT Braak stage-dependent differential expression. (a-f) Volcano plots showing -log₁₀(p-value) versus effect size for comparisons of mass-spectrometry-derived protein intensities. Panels a–c represent inferior temporal (IT) cortex (Mid vs. Low, High vs. Low, High vs. Mid), and panels d–f represent middle frontal (MF) cortex with the same comparisons. Significant proteins are shown as red (increased) or blue (decreased) in the higher Braak group, while non-significant proteins are shown in grey. Protein intensities were log₂-transformed and z-normalized. Effect sizes were estimated using linear mixed models with Braak stage and fraction as fixed effects. Proteins were considered significant at p < 0.005 with an absolute effect size ≥ 30%. Selected significant proteins are labeled. (**g**) Gene Ontology (GO) enrichment analysis of biological processes for differentially expressed proteins (DEPs) stratified by trend. The x-axis represents the enrichment score. Point color indicates statistical significance, and point size reflects the number of proteins in each category. (**h**) Sankey plot linking IT-MF concordant DEPs to biological pathways.

In the IT cortex, we identified 21, 62, and 75 upregulated DEPs in the comparisons of Mid-Braak vs. Low-Braak, High-Braak vs. Low-Braak, and High-Braak vs. Mid-Braak, respectively (Fig. 2a-c). Correspondingly, 2, 21, and 167 proteins were found to be downregulated in these comparisons, respectively. A total of 287 DEPs were identified across all three comparisons, including 58 (20%) known MAPT binding proteins and 51 (17%) APP/Aβ binding proteins. Top significant upregulated DEPs included MAPT, heparin-binding growth factor MDK, autophagy protein SQSTM1, estrogen growth factor receptor EGFR, calcium binding protein SMOC1 which interacts with Aβ pathology and is considered one of the earliest indicators of preclinical AD [5], and reactive astrocyte markers VIM and GFAP. Top down-regulated DEPs included endosomal regulator MTMR9, membrane scaffolding protein MPP1, synaptic protein PPFIBP1, lipid kinase DGKA and PRKCE with known involvement in tau phosphorylation, all with most pronounced changes between Mid- and High-Braak.

In the MF cortex, 51, 12, and 84 DEPs were upregulated, while 58, 5, and 18 were downregulated in the comparisons of Mid-Braak vs. Low-Braak, High-Braak vs. Low-Braak, and High-Braak vs. Mid-Braak, respectively, resulting in a combined total of 195 DEPs (Fig. 2d-f). Among these, 45 (23%) were known MAPT-binding proteins and 32 (16%) were APP/Aβ-binding proteins. The most significantly upregulated DEPs included MAPT, complement components (C4A, C4B, C3, C1S), the autophagy adaptor SQSTM1, collagen COL25A1, and MDK which inhibits Aβ aggregation and plaque formation [56]. In contrast to the IT region, a greater number of proteins were downregulated from Low- to Mid-Braak, including the serine protease KLK6, ALDH7A1 (an aldehyde metabolism enzyme), ABCE1 (an ATP-binding cassette protein involved in translation regulation), RPS12 (a ribosomal protein), CCDC124 (a cell cycle–associated protein), and ADD3 (a cytoskeletal protein), among the most significant changes.

Across both regions, a total of 452 DEPs were identified, including 220 upregulated and 232 downregulated proteins, based on the comparison showing the most pronounced (lowest p value) differences (Supplementary Table S1). The top enriched biological processes among upregulated DEPs included positive regulation of synaptic transmission (n=9), regulation of leukocyte migration (n=11), intermediate filament cytoskeleton organization (n=6), negative regulation of hydrolase activity (n=15), and protein-containing complex assembly (n=15) (Fig. 2g). In contrast, downregulated DEPs were enriched in processes such as regulation of neuron projection development (n=19), organelle localization (n=16), purine nucleotide metabolism (n=16), and endoplasmic reticulum–Golgi vesicle-mediated transport (n=9) (Fig. 2g).

### Differential functional profiles of DEPs in MF and IT cortex

Among the identified DEPs, 24 proteins were significantly associated with Braak stage in both IT and MF for the same Braak stage comparison and were therefore considered high-confidence shared proteomic changes. Figure S2 illustrates the distribution of protein abundances across NFT Braak groups stratified by region and biochemical fraction. To identify additional proteins that may represent concordant Braak-associated changes but did not reach statistical significance in both tissues, we calculated fraction-adjusted correlations between IT and MF protein abundances. An additional 28 proteins exhibited strong inter-regional concordance (ρ > 0.6), suggesting that they may reflect broader Braak-associated proteomic alterations shared across brain regions. These concordant Braak-associated proteins included several proteins with established links to AD pathology, including MAPT, the core component of NFT; SMOC1, COL25A1, SFRP1, and MDK, which are associated with amyloid plaque-related extracellular matrix remodeling; and GFAP and S100A6, which reflect glial responses. Functional enrichment analysis revealed overrepresentation of proteins involved in signaling by receptor tyrosine kinases (n=6), regulation of cytoskeleton organization (n=7), negative regulation of immune system process (n=7), cellular response to stress (n=6), regulation of vesicle mediated transport (n=7), and autophagy (n=3) (Fig. 2h).

We performed Gene Ontology to compare the functional profiles of DEPs identified in the IT and MF regions (Fig. 3a-3b). For IT DEP, we observed significant enrichment of proteins involved in intermediate filament-based processes (n=11), neuron projection development (n=22), and phospholipid metabolism (n=18). At the cellular component level, they were enriched in the vesicle lumen (n=21), primary lysosome (n=12), intermediate filament cytoskeleton (n=15), and ficolin-1-rich granule (n=12). For molecular function, enrichments included cadherin binding (n=22), protein kinase C binding (n=7), and SNARE binding (n=8). In contrast, MF DEPs were enriched in proteins involved in response to metal ions (n=14), actin filament organization (n=16), protein localization to endosomes (n=4), and protein kinase A signaling (n=4). Cellular component enrichments included blood microparticles (n=10), intercellular bridges (n=9), and microtubules (n=19). For molecular function, MF DEPs were enriched in guanyl nucleotide binding (n=16), phosphatase binding (n=7), and GTPase activity (n=12).

**Fig. 3.**
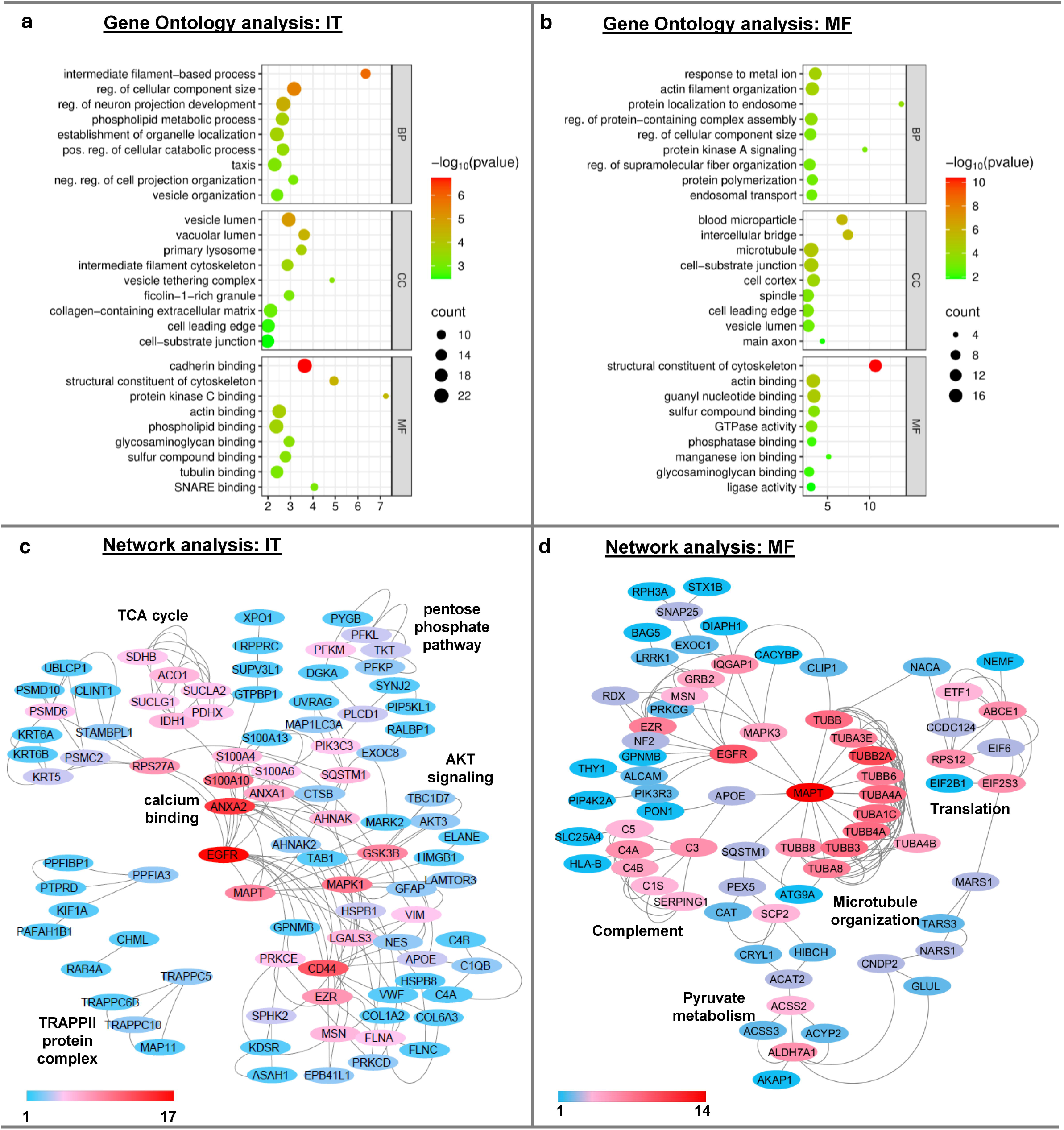
Functional enrichment analysis of NFT Braak stage-dependent differentially expressed proteins (DEPs). Panels **a–b** illustrate Gene Ontology (GO) enrichment analysis of DEPs identified in the inferior temporal cortex (**a**) and middle frontal cortex (**b**). The x-axis represents the enrichment score; point color denotes statistical significance, and point size indicates the number of proteins enriched in each GO term (BP, biological process; MF, molecular function; CC, cellular component). Panels **c–d** depict protein–protein interaction network analysis of DEPs generated using STRING-db and visualized in Cytoscape for inferior temporal cortex (**c**) and middle frontal cortex (**d**). Node color reflects connectivity, with red indicating a higher number of connections and blue denoting a lower number of connections.

Network analysis also revealed regional differences in key functional clusters among the DEPs (Fig. 3c-d). For IT DEPs, the enriched pathway clusters included calcium binding, the pentose phosphate pathway, the citrate cycle, AKT signaling, and the TRAPP complex. For MF DEPs, network analysis identified enrichment in microtubule cytoskeleton organization, complement activation, translational termination, and regulation of protein localization to the early endosome and pyruvate metabolism.

### Bidirectional abundance changes highlight greater dynamic variability in MF regions

In both IT and MF regions, we observed more DEPs when comparing Mid-Braak to High-Braak stages than when comparing Low-Braak to High-Braak. This suggests dynamic, non-monotonic changes in protein abundance during NFT progression, since a monotonic pattern would be expected to produce the largest differences between Low- and High-Braak stages. To identify bidirectionally expressed proteins (BEPs), Braak NFT stage group was modeled as an ordinal variable in the linear mixed-effects models, including both linear and quadratic terms. As shown in Fig. 4a-b, many proteins (164 in MF and 90 in IT) exhibited a significant quadratic term (p < 0.005 and |β| > 0.3), indicating a non-linear association with Braak NFT stage. To exclude proteins with trivial or noise-level fluctuations, we defined BEPs as those with significant quadratic effects and displaying significant pairwise differences between Mid-Braak and Low-Braak, as well as between High-Braak and Mid-Braak (p < 0.05). Interestingly, although the IT region contained more DEPs overall, a substantially larger number of BEPs were identified in the MF region (n=141) compared with IT (n=42; Supplementary Table S3). The Wnt signaling modulator Dickkopf-3 (DKK3) reported to be involved in Aβ clearance [52], was the only protein shared between the two cortical regions. In addition, seven proteins, RTN3, RPH3A, PIN1, OMG, ARHGAP26, FXR2 and TKT were among BEPs that exhibited strong inter-tissue correlations.

**Fig. 4.**
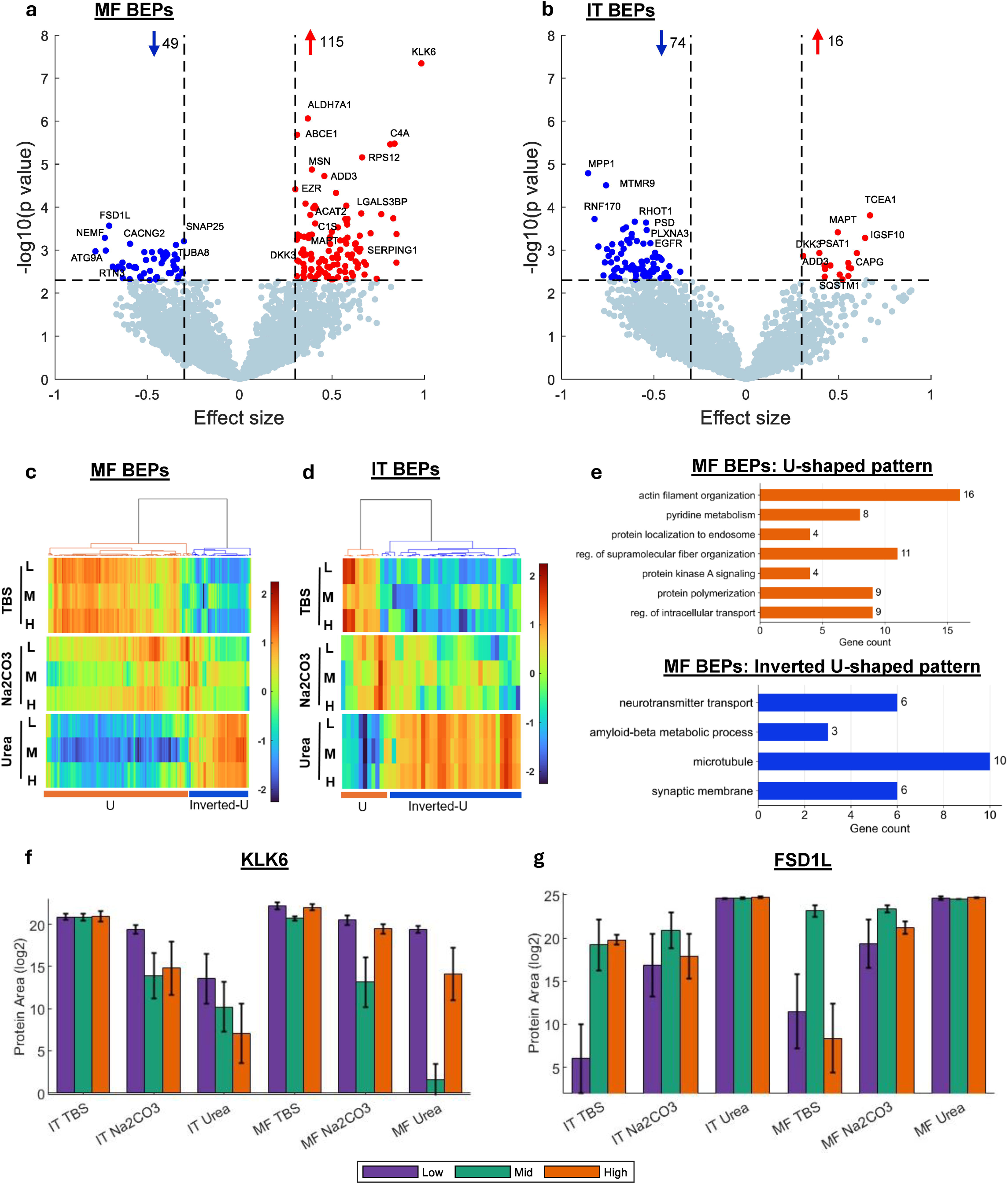
Proteins with bidirectional NFT Braak stage-associated expression changes. (**a-b**) Volcano plots of -log₁₀(p-value) versus effect size in the middle temporal cortex (MF; **a**) and inferior frontal cortex (IT; **b**). Red dots indicate proteins with U-shaped changes (down–up), and blue dots indicate inverted U-shaped changes (up–down). Non-significant proteins are shown in grey. Protein intensities were log₂-transformed and z-normalized. Effect sizes were derived from linear mixed models including Braak stage (linear and quadratic terms) and fraction as fixed effects. Proteins were considered significant at p < 0.005 with an absolute effect size ≥ 30%. Selected proteins are labeled. (**c–d**) Heatmaps show clusters of bidirectionally expressed proteins (BEPs) in MF (c) and IT (d), grouped by their expression patterns (U-shaped versus inverted U-shaped). The heatmaps were generated using mean log₂-transformed, z-normalized protein intensities stratified by fraction, NFT Braak stage, and tissue type. Clustering was performed using Euclidean distance with Ward’s linkage. (**e**) Functional enrichment analysis of MF BEPs by trajectory. (**f-g**) Bar plots showing log₂-transformed peak areas of KLK6 (f) and FSD1L (g).

Fig. 4c–d show heatmaps of the mean protein area (z-normalized) of BEPs in MF and IT, respectively, revealing two distinct trajectory patterns across the three Braak stage groups: U-shaped (positive quadratic term; down–up) and inverted U-shaped (negative quadratic term; up–down). In the MF region, U-shaped trajectories predominated (97 of 141 BEPs), whereas inverted U-shaped patterns were less frequent (44 of 141) (Fig. 4c). In contrast, most IT BEPs exhibited inverted U-shaped trajectories (n = 33 of 42), with fewer showing U-shaped patterns (n = 9 of 42) (Fig. 4d).

We compared the functional profiles of MF BEPs exhibiting the two distinct trajectory patterns using DAVID Bioinformatics Tools (Fig. 4e). Notably, MF BEPs with U-shaped patterns were significantly enriched for actin-binding proteins (n = 11) and for pathways related to early-to-late endosome transport (n = 5), carbon metabolism (n = 8), the complement cascade (n = 6), and apoptosis (n = 6). Network analysis (Supplementary Fig. S3) further supported enrichment in cytoplasmic translation, complement activation, and protein localization to early endosomes. In contrast, BEPs with inverted U-shaped patterns were enriched for microtubule-associated proteins (n = 12) and proteins involved in the synaptic vesicle cycle (n = 4) and intracellular transport (n = 12). Network analysis indicated enrichment in processes related to microtubule-dependent trafficking from the Golgi to the plasma membrane and synaptic vesicle fusion at the presynaptic active zone. Among MF BEPs, the serine protease KLK6, an amyloid-related enzyme, was the most prominent protein exhibiting a U-shaped trajectory (Fig. 4f), whereas FSD1L (fibronectin type III and SPRY domain-containing 1-like protein) showed the strongest inverted U-shaped pattern (Fig. 4g).

Gene Ontology analysis did not reveal significant pathway enrichment for IT BEPs, likely reflecting the limited number of features; however, several inverted U-shaped IT BEPs (n = 15) are involved in catabolic processes. Notably, U-shaped IT BEPs included RAB11A and RAB11B, key regulators of recycling endosome trafficking that may influence Aβ clearance. The most prominent IT BEPs included the membrane-associated cytoskeleton-interacting scaffold protein MPP1, myotubularin-related protein 9 (MTMR9), transcription elongation factor A protein 1 (TCEA1), and the E3 ubiquitin-protein ligase RNF170.

### Fractional abundance shifts reveal the association of extensive protein conformational alterations with tau NFT pathology in both cortical regions

Protein changes may influence cellular phenotypes not only through changes in abundance but also through conformational alterations that affect their solubility. To investigate this, we applied linear mixed-effects models incorporating an interaction term between Braak NFT group and biochemical solubility fraction (TBS, Na_2_CO_3_, and Urea; treated as ordinal levels 1–3, as described in Methods) to identify differentially soluble proteins (DSPs). Proteins are considered more soluble if their proportion increases in the TBS fraction. Many proteins showed significant interaction effects (p < 0.005, |β| ≥ 0.3), indicating Braak NFT stage-dependent solubility changes (Fig. 5a–f).

**Fig. 5.** Identification of proteins with NFT Braak stage-associated differential solubility. (**a–f**) Volcano plots showing -log₁₀(p-value) versus effect size. Panels **a–c** show results for inferior temporal (IT) cortex (Mid vs. Low, High vs. Low, and High vs. Mid), and panels **d–f** show results for middle frontal (MF) cortex with the same comparisons. Proteins with decreased solubility at higher Braak stages are shown in red, those with increased solubility in blue, and non-significant proteins in grey. Protein intensities were log₂-transformed and z-normalized. Effect sizes were β coefficients for the Braak × fraction interaction term in linear mixed models. Only selected proteins are labeled. (**g**) Venn diagram showing the overlap of differentially soluble proteins (DSPs) identified in the IT and MF cortices. (**h**) Venn diagram showing the overlap among differentially expressed proteins (DEPs), bidirectionally expressed proteins (BEPs), and DSPs identified in this study. (**i**) Stack bar graphs illustrate the fractional distribution of selected DSPs stratified by tissue type and NFT Braak group. Error bars represent the standard error of the mean.

In both IT and MF regions, DSPs with negative interaction terms (greater solubility at higher Braak NFT stage) were mainly observed comparing Low-Braak and Mid-Braak groups (51 in IT, 59 in MF). In contrast, DSPs with positive interaction terms (lower solubility at higher Braak NFT stage) were predominantly identified comparing Mid-Braak and High-Braak groups (521 in IT, 128 in MF). These findings indicate differential dynamic regulation of protein solubility alongside abundance-based changes at early versus late stages of NFT progression in AD.

Adding the requirement of significant abundance differences in either TBS or Urea fractions (p < 0.05), we identified 693 DSPs, including 562 in the IT region and 185 in the MF region, with 54 shared (Fig. 5g and Supplementary Table S4). An additional 98 DSPs exhibited strong inter-tissue concordance, yielding a total of 152 concordant DSPs. These proteins were significantly enriched for chaperone (n=17) and cytoskeletal proteins (n=23), as well as proteins involved in mRNA processing (n=11), autophagy regulation (n=12), and regulation of synapse organization (n=12), highlighting common disruptions in proteostasis, cellular architecture, and neuronal function associated with NFT progression. Among all DSPs, 172 were also DEPs or BEPs (Fig. 5h). Notably, 156 (22.5%) DSPs are known tau interactors, and 121 (17.5%) bind APP/Aβ. As expected, MAPT was the most significant DSP in both regions. Other top-ranked proteins included the spliceosome component SNRNP70, lipoprotein APOE, and the autophagy adaptor SQSTM1 (Fig. 5i). MAPT, SNRNP70, and APOE had higher proportions present in the Urea fraction in the high-Braak group, indicating lower solubility (Fig. 5h). In contrast, SQSTM1 showed the opposite pattern, with high-Braak NFT stage samples displaying a greater proportion in the TBS fraction, indicating greater solubility (Fig. 5h).

Across all six comparisons, 75 DSPs showed increased solubility (32 in IT and 43 in MF) and 572 showed decreased solubility with higher Braak NFT stage (496 in IT and 114 in MF, with 38 overlapped; DSPs with mixed trends in IT and MF were not included in the count; Supplementary Table S3). DSPs with increased solubility were significantly enriched for synaptic proteins (n=17) and endosome-associated proteins (n=12) and included several with known associations with neurodegeneration (DNAJC13, FGGY, L1CAM, GRN, KIF1C, SQSTM1, and UBA5). As shown in Supplementary Fig. S4a, DSPs with decreased solubility were significantly enriched in biological processes such as protein-RNA complex organization (n=28), endosomal transport (n=34), actin filament organization (n=44), protein folding (n=25), and synapse organization (n=40). Enriched cellular components included glutamatergic synapse (n=36), and early endosome (n=36), cell-substrate junction (n=38). Molecular function enrichment included tau protein binding (n=10), actin binding (n=37), heat shock protein binding (n=15), and phosphoprotein binding (n=13) and coated membrane (n=18). Supplementary Fig. S4b-g illustrate protein–protein interaction networks based on STRING-DB analysis, revealing significant clustering of DSPs within networks related to ubiquitin-proteasome complex (n=23), mRNA splicing via the spliceosome (n=18), translation elongation (n=13), protein neddylation (n=12), chaperone complexes (n=10), and the retromer/WASH complex (n=9).

## Discussion

Mass spectrometry–based proteomics is increasingly used to profile large numbers of proteins for investigations into the molecular mechanisms underlying human disease owing to substantial advances in proteomic technologies as well as computational analysis capabilities [41, 43]. Here, we applied state-of-the-art nanoLC–MS/MS for unbiased proteomic profiling of two neocortical brain regions affected differentially by tau pathology. Integration of LC–MS/MS with solubility-based biochemical fractionation enabled quantification of 5,573 proteins and assessment of their associations with NFT neuropathology staging, capturing spatial, temporal, and conformational protein alterations linked to tau molecular biology. Across analyses, numerous proteins exhibited Braak NFT stage–associated differential expression patterns (DEP: 287 in IT; 195 in MF), dynamic Braak-dependent patterns referred to as bidirectional expression patterns (BEP: 42 in IT; 141 in MF), or differential solubility patterns (DSP: 562 in IT; 185 in MF), collectively identifying 1,012 proteins with significant tau NFT-related alterations.

Functional analysis of the 452 DEPs identified in our study revealed enrichment of pathways consistent with tau related pathobiology, including cytoskeletal remodeling, proteolytic and inflammatory processes, as well as neuronal structure, intracellular trafficking, metabolism, and synaptic maintenance—processes broadly implicated in AD pathogenesis [7, 24, 28]. Notably, 359 (79.4%) of these DEPs were reported in at least one of the 38 human AD brain proteomic studies compiled by Askenazi et al., with 120 (26.5%) included in the subset of 848 proteins consistently altered across at least five studies [4], indicating strong concordance with prior findings. These proteins prominently map to established AD-related pathways such as neuroinflammation (e.g., complement components C1–C5 and SERPING1), synaptic function (e.g., SNAP25, STX1B), cytoskeletal organization (e.g., GFAP, VIM, PLEC), actin cytoskeleton dynamics (e.g., CAPG), endolysosomal trafficking (e.g., VPS41, RAB4, RAB4B), and mitochondrial metabolism (e.g., SUCLA2, PDHX), and autophagic process (ATG9A, MAP1LC3A, SQSTM1, CTSB).

A number of proteins identified in our study were not among the subset of DEPs reported in the Askenazi et al. compilation but are nonetheless linked to AD-relevant biological processes, including cytoskeletal organization and vesicular trafficking (e.g., ARHGAP family, TRAPPC complexes, WDR81), mitochondrial metabolism (e.g., MT-CO1, COQ5), RNA regulatory pathways (e.g., ABCE1, MTREX, PRPF4), proteostasis (e.g., PSMD10, UBE2F), and lipid metabolism (e.g., SCP2, SPTLC2). Together, these findings highlight the robustness of MS-based proteomics in capturing both well-established and underexplored molecular alterations associated with AD pathology, supporting our hypothesis.

Several observations from our study support the idea that the IT and MF regions are differentially vulnerable to tau pathology and/or respond differently to its development, including the results of differential regional patterns of DEPs in relation to tau pathology progression. IT and MF were similar in showing more DEPs upregulated with greater NFT progression, but they differed in numbers of downregulated DEPs: in IT more DEPs were downregulated with more advanced NFT progression (i.e., Braak stage III/IV to Braak stage V/VI groups), while in MF more DEPs were downregulated early (i.e., Braak stage 0-II to Braak stage III-IV).

Combining shared DEPs and those with strong inter-tissue correlation, we identified 52 concordant DEPs between the two regions, and they may represent more generalized (but limited) protein change relevant to tau pathology. Notably, several concordant DEPs have established links to AD-related pathological processes, including core tau pathology (MAPT), plaque-associated and extracellular matrix remodeling (COL25A1, SMOC1, MDK), innate immune activation and neuroinflammation (C4A, C4B, GPNMB, NLRX1), lysosomal and autophagic pathways (SQSTM1, PPT1, TPP1, PITRM1), oxidative stress responses (PRDX1, PRDX6, FTL), cytoskeletal organization and cellular remodeling (MSN, ARHGAP26, CAPG, ARPC1B), and synaptic function and vesicular trafficking (RAB4A, RPH3A, VGF, KIF1A). The prominent involvement of shared DEPs in AD-related processes suggests that these proteins may represent core molecular mechanisms that drive disease progression across brain regions.

Distinct functional profiles of IT DEPs versus MF DEPs further support differential regional responses to tau pathology. Interestingly, while both regions showed alterations in cytoskeletal organization, IT DEPs were enriched in intermediate filament processes, whereas MF DEPs were enriched in actin and microtubule-related pathways. These cytoskeletal systems have distinct properties yet collectively support the structural and functional plasticity of neuronal architecture [8, 21, 28, 31]. Actin filaments and microtubules are more dynamic and play essential roles in synaptic plasticity and intracellular transport, whereas intermediate filaments provide structural stability. These findings suggest that protein changes in MF, such as active cytoskeletal remodeling, may be related to the area’s reported plasticity response to AD pathology. Consistent with this, several actin/microtubule-associated proteins exhibited bidirectional expression changes, increasing from low to mid Braak stages and decreasing thereafter. As tau pathology promotes microtubule instability, the early upregulation of actin and microtubule components may represent a compensatory response to maintain cytoskeletal integrity and intracellular transport. One the other hand, the enrichment of intermediate filament processes in IT may indicate attempts at structural stabilization prior to NFT maturation and cellular dysfunction.

In addition to those mentioned above, substantial number of additional proteins exhibited dynamic, rather than monotonic, changes across Braak stages. In total, 183 BEPs were identified based on multiple selection criteria, including a significant quadratic association with Braak stage, as well as significant differences between Low- and Mid-Braak and between Mid- and High-Braak stages. The majority of BEPs (144/183) were also identified as DEPs; however, their largest changes were observed between adjacent stages (Low-Braak vs Mid-Braak or Mid-Braak vs High-Braak) rather than between Low-Braak and High-Braak stages, as would be expected under a monotonic pattern. Our findings differ from meta-analysis by Askenazi et al., which found that most proteins (240 of 258) exhibiting significant changes in early and advanced AD followed a consistent directional trend [4]. This discrepancy may reflect differences in study design, as the Askenazi et al. staging of AD was clinically (not neuropathologically) based, with their “early-stage” AD referring to MCI or pre-clinical AD cases and “advanced stage” referring to a typical AD. Further, they integrated data across multiple cohorts, whereas our observations are derived from a single cohort. In addition, the studies included in Askenazi et al. primarily relied on pairwise group comparisons without intermediate groups, which may not be able to capture non-monotonic, stage-dependent changes.

The observed bidirectional changes are consistent with the increasingly recognized dynamic and context-dependent regulation of important biological processes during AD progression such as neuroinflammation and its effects on synapses. For example, neuroinflammation, largely mediated by reactive astrocytes and microglia, is now considered a double-edged sword, contributing to early protective functions such as Aβ clearance and synaptic remodeling, but transitioning toward sustained activation with toxic effects that promote synaptic loss and neurodegeneration at later stages [29, 32]. Supporting this concept, Sun et al. identified multiple distinct microglial states from ∼194,000 single-nucleus transcriptomes and epigenomes across human brains, demonstrating that microglia undergo stage-dependent transitions during AD progression [48]. Similarly, complement-mediated synaptic pruning, which is essential for physiological circuit refinement, becomes aberrantly reactivated and contributes to pathological synapse loss in AD [34].

We observed substantially more BEPs in MF than in IT (141 vs. 42). This may reflect more dynamic and transitional protein changes during early tau pathology development in the MF region at the time when NFT load is already substantial in the IT region which shows more cumulative and monotonic alterations, in concert with the idea of differential vulnerability and response to tau pathology of the two regions. The minimal overlap between regions (only one shared BEP) further indicates region-specific responses to tau pathology. MF BEPs with inverted U-shaped pattern, enriched in microtubule-associated, synaptic, and transport pathways, likely represent early compensatory upregulation of neuronal function that subsequently deteriorates. Conversely, U-shaped MF BEPs, enriched in actin remodeling, endosomal trafficking, metabolism, and complement activation, may indicate delayed activation of stress, inflammatory, and compensatory pathways as pathology advances.

KLK6, a CNS-enriched serine protease with multifaceted roles in neurodegeneration, including clearance of Aβ and tau aggregate, synaptic function, and neuroinflammatory signaling [9, 33], exhibited one of the most pronounced bidirectional changes in MF, but a more monotonic pattern in IT, particularly in the Na₂CO₃ and Urea fractions. Although previous studies have reported altered KLK6 levels, it was not identified as a DEP in the datasets compiled by Askenazi and Yarbro [3, 4, 16, 53]. Our results suggest that KLK6 dysregulation is dynamic and region-dependent, which may contribute to this inconsistency. Altered KLK6 in Na₂CO₃ (membrane-enriched) and Urea (aggregate-enriched) fractions, with minimum change in the TBS pool, suggest that its influence on AD progression may be through interaction with membrane-associated and aggregated pathological substrates.

Another notable finding of our study is the large number of proteins exhibiting Braak NFT stage-associated changes in solubility. Specifically, DSPs showing greater solubility with more tau pathology were mainly observed early (Low-vs-Mid Braak stage), similarly in both regions (51 in IT, 59 in MF), suggesting that both regions initially resist tau pathology development. However, DSPs showing lower solubility at higher Braak NFT stage were mainly seen at the advanced tau pathology (Mid- vs. High-Braak stage) with greater abundance in IT cortex (521 in IT, 128 in MF) suggesting higher vulnerability of the IT region to protein aggregation and/or an MF compensatory plasticity response that resists protein aggregation. While lower solubility of proteins with more advanced Braak NFT stage may partly reflect their sequestration into Aβ and tau aggregates, functional enrichment analysis suggests selective involvement of proteins central to proteostasis, including the translational machinery, ubiquitin–proteasome system, molecular chaperones, and endosomal pathways, highlighting disruption of protein quality control and degradation mechanisms during disease progression and adding further support to the concept of differential regional vulnerability in AD.

This study has several limitations. First, given the large number of proteins analyzed relative to the available sample size, we didn’t perform multiple comparison correction which would substantially reduce statistical power. Although a combination of a more stringent significance threshold (p < 0.005) and a beta estimate cutoff was used to reduce the inclusion of marginal changes, the reported associations should be considered exploratory. In addition, biological interpretation was primarily based on pathway-level analyses using GO enrichment, and the identified proteomic changes were not validated using independent orthogonal approaches. Therefore, our findings should be regarded as hypothesis-generating and require independent replication and validation in larger cohorts. Second, our study relied on LC-MS/MS-based proteomic measurements, and the depth and coverage of the proteomic data may be influenced by variability in protein extraction efficiency, enzymatic digestion, chromatographic retention, and ionization efficiency. In addition, protein abundance estimates derived from peptide-centric shotgun proteomics have inherent limitations that may affect quantitative accuracy. Third, although amyloid-β and tau pathologies are both defining features of AD, our analyses primarily focused on NFT Braak stage as an index of disease severity. Due to the strong correlation between amyloid and tau pathology in our cohort, it was not possible to reliably distinguish their individual contributions to the observed proteomic changes. Consequently, the identified proteins should be interpreted as markers of progressive AD pathology rather than as tau-specific molecular alterations. Fourth, as this study is based on an autopsy cohort, longitudinal assessment within individuals is not feasible. While we used standard neuropathological assessment of tau pathology changes (Braak NFT Stages), the inferred stage-dependent disease progression represents a pseudo-trajectory and should be interpreted with caution. Finally, orthogonal approaches, such as immunohistochemistry assays, will help validate the spatial and biochemical alterations observed.

## Conclusions

Together, our study provides a systems-level view of proteomic changes associated with tau NFT pathology by integrating regional and neuropathology stage-dependent analyses. Our findings point to coordinated alterations spanning cytoskeletal organization, metabolic pathways, vesicular trafficking, and proteostasis, incorporating both dynamic, neuropathology stage-dependent changes in protein abundance and shifts in protein solubility. These results suggest that early AD pathology stages may involve region-specific dynamic and potentially adaptive responses that evolve into more cumulative changes at later stages. Importantly, the regional and temporal heterogeneity observed, together with dynamic and solubility-associated protein alterations, highlights the complexity of AD pathobiology and the challenges in translating brain tissue-derived findings into diagnostic biomarker discovery.

## Abbreviations

Aβ: Amyloid-beta
AD: Alzheimer’s disease
AGC: Automatic gain control
BEP: Bidirectionally expressed protein
DEP: Differentially expressed protein
DSP: Differentially soluble protein
DTT: Dithiothreitol
FA: Formic acid
IAA: Iodoacetamide
IT: Inferior temporal cortex
LC-MS/MS: Liquid chromatography-tandem mass spectrometry
MF: Middle frontal cortex
MS: Mass spectrometry
NFT: Neurofibrillary tangle
PCA: Principle component analysis
PET: Positron emission tomography
PI: Protease inhibitor
SDS: Sodium dodecyl sulfate
SP3: Single-pot, solid-phase-enhanced sample-preparation
SPE: Solid phase extraction

## Acknowledgments

We are indebted to the participants, family members, and staff of the Pitt-ADRC study. The content of the manuscript is solely the responsibility of the authors and does not necessarily represent the official views of the Department of Veterans Affairs or the United States Government.

## Author contributions

TKK, MDI, and XZ contributed to the study’s conception and design. XZ, WRP, and VNP performed the experiments. JK, MDI, and EEA contributed to neuropathological data collection and analysis. XZ performed mass spectrometry analysis, data analysis and produced the figures. XZ, EEA, WGB, MDI, and TKK were major contributors in writing the manuscript. All authors contributed to and approved the final version of the manuscript.

## Data availability

De-identified, cohort-level data will be shared at the request of verified investigators to replicate procedures and results reported in this article. Data transfer agreements in accordance with US legislation and the decisions of the University of Pittsburgh’s Institutional Review Board, may need to be established.

## Declarations

### Ethics approval and consent to participate

All brain tissue samples were obtained with full written informed consent and approved by the Committee for Oversight of Research and Clinical Training Involving Decedents (CORID) and the University of Pittsburgh Institutional Review Board.

### Consent for publication

Not applicable.

