## Supplementary figures 1-4 for "Brain region- and solubility-dependent proteomic signatures of neurofibrillary tangle pathology in Alzheimer’s disease"

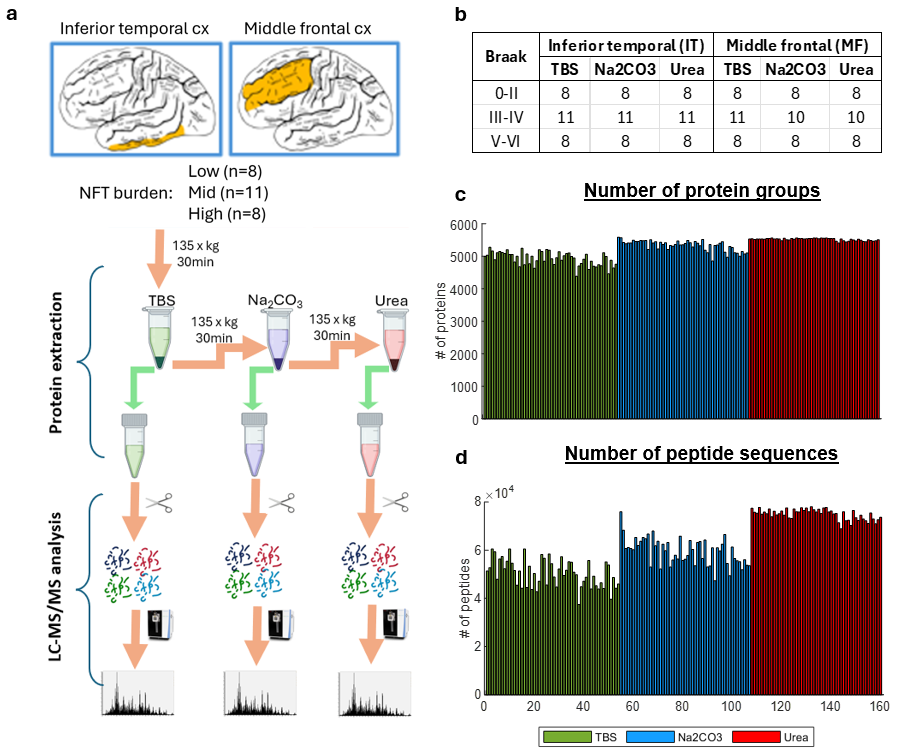

**Figure S1: Overview of experimental design and proteomic profiling workflow.** (**a**) Schematic illustration of the experimental workflow. Inferior temporal and middle frontal cortices from 27 post‑mortem brains were sequentially fractionated into TBS‑soluble, Na₂CO₃‑soluble, and urea‑soluble fractions, followed by trypsin digestion and LC–MS/MS analysis. A total of 5,573 protein groups were identified with a minimum of two unique peptides. (**b**) Overview of brain homogenate samples included in the study. (**c**) Bar graphs showing the number of proteins identified in each sample. (**d**) Bar graphs showing the number of peptide sequences identified in each sample.

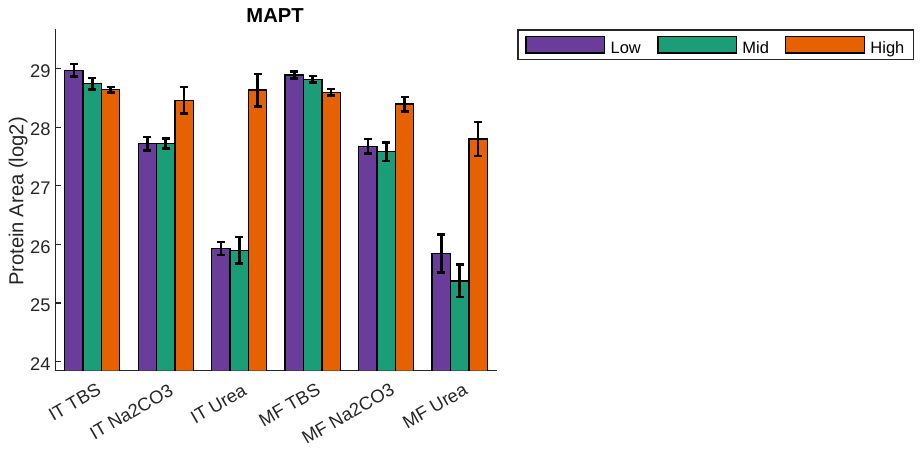

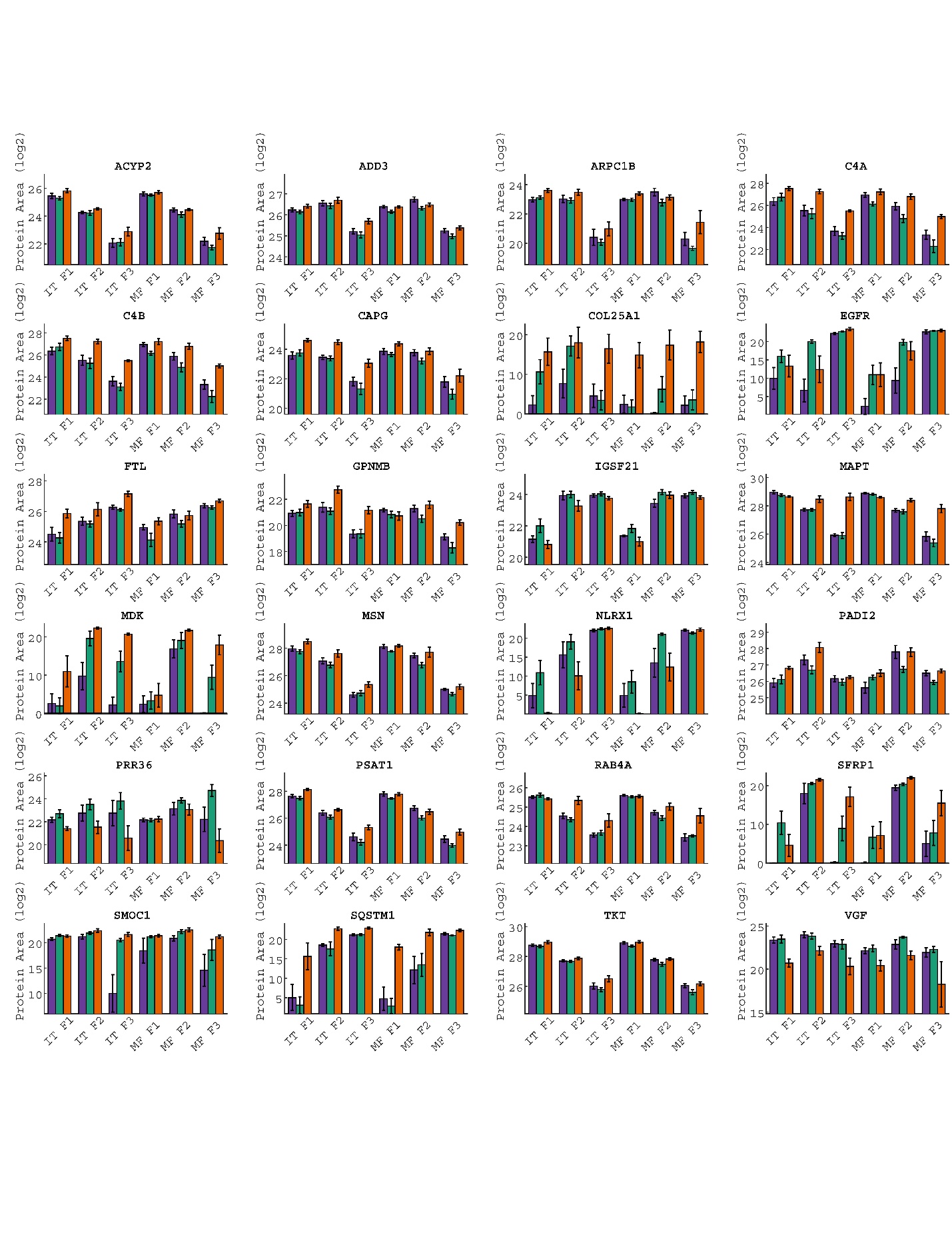

**Figure S2:** **Bar plots of differentially expressed proteins (DEPs) shared between inferior temporal and middle frontal cortices.** The y-axis represents log₂-transformed protein area normalized to the total protein area of each sample. Error bars indicate the standard error of the mean. Low, Mid, and High correspond to NFT Braak stages 0–II, III–IV, and V–VI, respectively. p < 0.05; p < 0.01; p < 0.001. F1, F2, and F3 denote the TBS‑soluble, Na₂CO₃‑soluble, and urea‑soluble fractions, respectively.

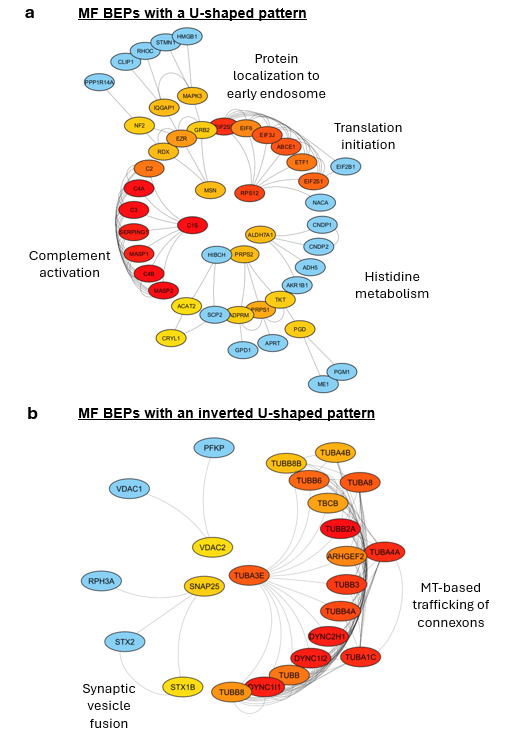

**Figure S3:** **Protein–protein interaction network analysis of bidirectionally expressed proteins (BEPs) in the middle frontal cortex.** Panel **a** and **b** are for BEPs with U‑shaped (decrease followed by increase; a) or inverted U‑shaped (increase followed by decrease; b) expression patterns, respectively. Networks were generated using STRING‑db and visualized in Cytoscape. Node color reflects connectivity, with red indicating higher connectivity and blue indicating lower connectivity.

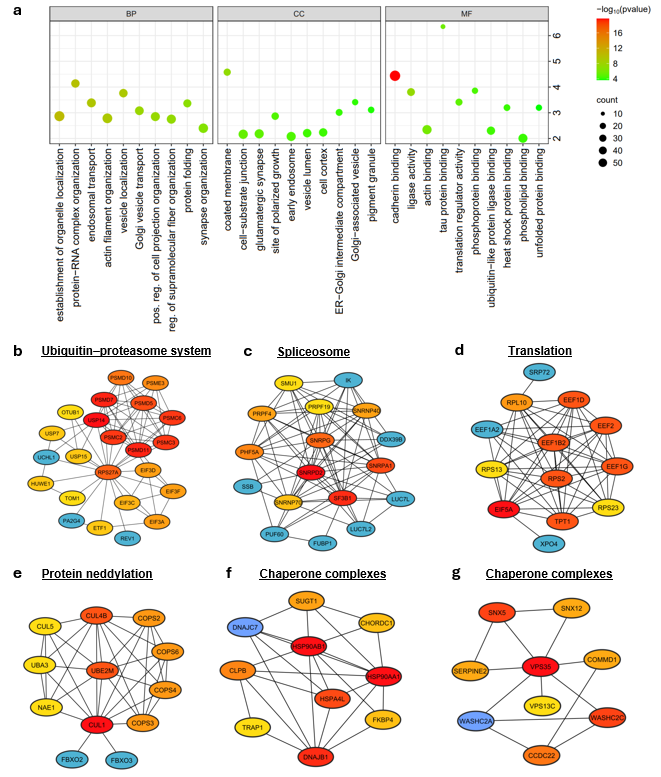

**Figure S4:** **Functional enrichment analysis for proteins with NFT Braak stage-dependent differential solubility.** Panels **a** illustrates Gene Ontology (GO) enrichment analysis. The enrichment score is shown in the y‑axis. Point color denotes statistical significance, and point size indicates the number of proteins enriched in each GO term (BP, biological process; MF, molecular function; CC, cellular component). Panels **b–g** show protein-protein interaction network analyses of differentially soluble proteins generated using STRING‑db and visualized in Cytoscape. The networks represent UCH proteases and the proteasome complex (**b**), mRNA splicing via the spliceosome (**c**), translation elongation (**d**), protein neddylation (**e**), chaperone complexes (**f**), and the retromer/WASH complex (**g**). Node color reflects connectivity, with red indicating a higher number of connections and blue indicating a lower number of connections.
